# Structural network topology mediated cognitive impairment in Parkinson’s disease

**DOI:** 10.1101/2023.09.23.23295998

**Authors:** Zhichun Chen, Guanglu Li, Liche Zhou, Lina Zhang, Li Yang, Jun Liu

## Abstract

Cognitive impairment (CI) is one of the most prominent non-motor symptoms in Parkinson’s disease (PD). How brain network abnormalities contribute to CI in PD patients remain largely unclear. The goal of this study is to explore whether aberrations of brain network topology were causally associated with cognitive decline in PD patients. PD patients receiving magnetic resonance imaging from Parkinson’s Progression Markers Initiative (PPMI) database were specifically selected. According to the scores of Montreal Cognitive Assessment (MoCA), PD patients were classified into CI+ group (MoCA score ≤ 25) and CI-group (MoCA score > 25) to investigate whether clinical features and brain networks were significantly different between two groups. Mediation analysis was utilized to evaluate whether brain network alterations contributed to CI in PD patients. We revealed CI + group exhibited more severe non-motor symptoms compared to CI-group. In addition, age, excessive daytime sleepiness, and depressive symptoms were found to be significantly associated with CI of PD patients. Moreover, CI+ group exhibited statistically different local topological properties in structural network compared to CI-group. Furthermore, differential local topological metrics in structural network meditated the effects of age, excessive daytime sleepiness, and depression on cognitive decline of PD patients. Taken together, out study suggested that PD patients with CI exhibited notable disturbances of structural network topology, which mediated negative associations between of age, excessive daytime sleepiness, depression and cognitive decline of PD patients.

## 1. Introduction

Cognitive impairment (CI) is one of the most common non-motor manifestations in Parkinson’s disease (PD) [1] and significantly associated with disease progression and future prognosis [1–3]. Cognitive decline may occur at any disease stage, even at the early stage or the prodromal phase [1, 4–6]. Generally, PD patients can exhibit global cognitive decline and also specific impairment of cognitive domains, such as executive function [7, 8], verbal memory [9–11], episodic memory [12–14], visuospatial capacity [15, 16], and processing speed [17, 18]. Previous studies have shown that age, education, baseline cognition, hypertension, motor symptoms, depression, excessive daytime sleepiness (EDS), and anxiety were associated with cognitive decline in PD patients [19–24]. Consistently, our recent findings also showed that age was significantly associated with scores of multiple cognitive assessments in PD patients, including Symbol Digit Modalities Test (SDMT), Montreal Cognitive Assessment (MoCA), Benton Judgement of Line Orientation Test (BJLOT), Letter Number Sequencing Test (LNS), and Semantic Fluency Test (SFT), therefore, age is a cardinal demographic factor remarkably shaping cognitive ability in PD patients [25]. Interestingly, in a recent study, we also revealed that EDS was negatively associated with scores of BJLOT, SDMT, and MoCA [26]. Taken together, previous studies and our recent findings demonstrated that CI in PD patients was associated with multiple clinical features, including age and EDS. However, the specific neural mechanisms underlying the associations between these clinical features and cognitive impairment in PD patients remain largely unknown.

Functional magnetic resonance imaging (fMRI) is a wonderful tool to explore and evaluate how the functional and structural brain changes contributed to the onset and progression of neuropsychiatric diseases, including bipolar disorder, schizophrenia, PD, and Alzheimer’s disease (AD) [27–31]. In addition, fMRI has also been widely utilized to decipher the functional and structural abnormalities causally associated with cognitive deficits in PD patients [32–36]. Brain network analysis was an important neuroimaging approach [37–41] and has revealed widespread changes of structural and functional networks in PD patients with CI [34, 42–44]. Consistently, aberrations in both structural networks and functional network were found to be correlated with cognitive decline in PD patients [25, 34, 45]. For structural network, we recently revealed that small-world topology in structural network was significantly associated with verbal memory maintenance of PD patients [9]. Moreover, we also showed that global network metrics, such as global efficiency and local efficiency in structural network, partially mediated the age-dependent CI in PD patients [25]. For functional network, it has been shown that increased functional connectivity between left posterior cingulate cortex and left parahippocampal gyrus in default mode network (DMN) was associated with early cognitive decline in PD patients [42]. In addition, functional connectivity between left and right hippocampus in DMN was also associated with global and domain-specific cognitive decline in PD patients [34]. In agreement with these findings, the critical role of DMN in cognitive decline of PD patients has been supported by several previous studies [45–47]. Apart from DMN, other resting-state networks, such as frontostriatal network [33], dorsal attention network [32], frontoparietal network [32], and salience network [48], were also found to be associated with CI in PD patients. Taken together, both structural and functional network measurements contributed to CI in PD patients.

As we mentioned above, multiple clinical characteristics, such as age, EDS, and depression, were associated with CI [22–25], however, the specific network mediators underlying the negative associations between these clinical characteristics and CI remained elusive in PD patients. Recently, we revealed global topology in structural network mediated the effects of age on cognitive decline in PD patients [25], whether local topology in structural network also mediated the associations between above clinical characteristics and CI in PD was tremendously unclear. In a recent study, we demonstrated local topology in structural network mediated the effects of age on LNS, BJLOT, and SDMT scores in PD patients [49], however, whether local topology in structural network also mediated the effects of age or other clinical characteristics (i.e., EDS) on MoCA scores were poorly understood. To identify the local structural network mediators associated with CI, PD patients (n = 145) were divided into CI+ group (MoCA score < 26) and CI-group (MoCA score ≥ 26) according to the Movement Disorder Society PD Mild Cognitive Impairment (MDS PD-MCI) criteria [50, 51]. As a consequence, the goal of current study is to investigate whether age, EDS, and depression shape cognitive decline by influencing local structural network topology of PD patients. Specifically, our objectives include: (i) to compare the clinical manifestations between CI- and CI+ patients; (ii) to validate whether age, EDS, and depression are associated with CI of PD patients; (iii) to compare the local topology of structural network between CI- and CI+ patients; (iv) to explore the associations between local topological measurements and CI, age, EDS, and depressive symptoms; (v) to investigate whether differential local topological metrics mediate the effects of age, EDS, and depression on CI of PD patients.

## 2. Materials and Methods

### 2.1. Participants

A total of 145 PD patients from Parkinson Progression Markers Initiative (PPMI) database [52, 53] were included for analyses in the present study. The PPMI study was approved by the Ethics Committee of all participating sites and all participants signed informed consent prior to participation. The inclusion criteria for PD patients were as follows: (i) The participants were over 30 years of age; (ii) The participants met the MDS Clinical Diagnostic Criteria for PD; (iii) The participants received the evaluation of global cognition using MoCA; (iv) The participants received T1-weighted magnetization-prepared rapid acquisition gradient echo (MPRAGE) MRI and diffusion tensor imaging (DTI) simultaneously. The exclusion criteria for PD patients were as follows: (i) The participants showed obvious abnormalities in regular T1-weighted and T2-weighted MRI; (ii) The participants were diagnosed with AD or other types of dementia except PD dementia; (iii) The participants were diagnosed with systemic diseases that cause cognitive decline, including cardiovascular, liver, kidney, or endocrine diseases; (iv) The participants had severe mental illness, such as schizophrenia and depression; (v) The participants were diagnosed with other central nervous system diseases associated with cognitive decline, such as Wernicke encephalopathy, traumatic brain injury, prion disease, and glioma. (vi) The patients were prescribed with medications that may cause cognitive decline, such as antiepileptic or antipsychotic drugs. In line with above inclusion and exclusion criteria, a total of 145 PD participants were included for the final analysis. Each included participant underwent a battery of neuropsychological examinations. The motor assessments included Hoehn and Yahr (H&Y) stages, Tremor scores, Total Rigidity scores, and the MDS Unified Parkinson’s Disease Rating Scale (UPDRS) scores (part III and total scores). The non-motor assessments included Epworth Sleepiness Scale (ESS), REM Sleep Behavior Disorder Screening Questionnaire (RBDSQ), Scale for Outcomes in Parkinson’s Disease-Autonomic (SCOPA-AUT), 15-item Geriatric Depression Scale (GDS), State-Trait Anxiety Inventory (STAI), Hopkins Verbal Learning Test–Revised (HVLT-R: Immediate Recall and Delayed Recall), BJLOT, LNS, SFT, SDMT, and MoCA. The patients also obtained [^123^I] FP-CIT SPECT scans, which were examined in accordance with the technical manual of the PPMI study (http://ppmi-info.org/). The striatal binding ratios (SBRs) for bilateral caudate, putamen, and striatum were derived from SPECT scans. Specifically, the SBRs were calculated with the formula: (target region/reference region)-1, in which occipital lobe was the reference region. To examine the effects of CI on clinical assessments and local network topology, PD patients (n = 145) were classified into CI+ group (MoCA score < 26) and CI-group (MoCA score ≥ 26) according to the MDS PD-MCI criteria [50, 51], which entailed MoCA scores < 26. The clinical features of participants in each group were shown in Table 1 and Figure 1.

**Fig. 1.**
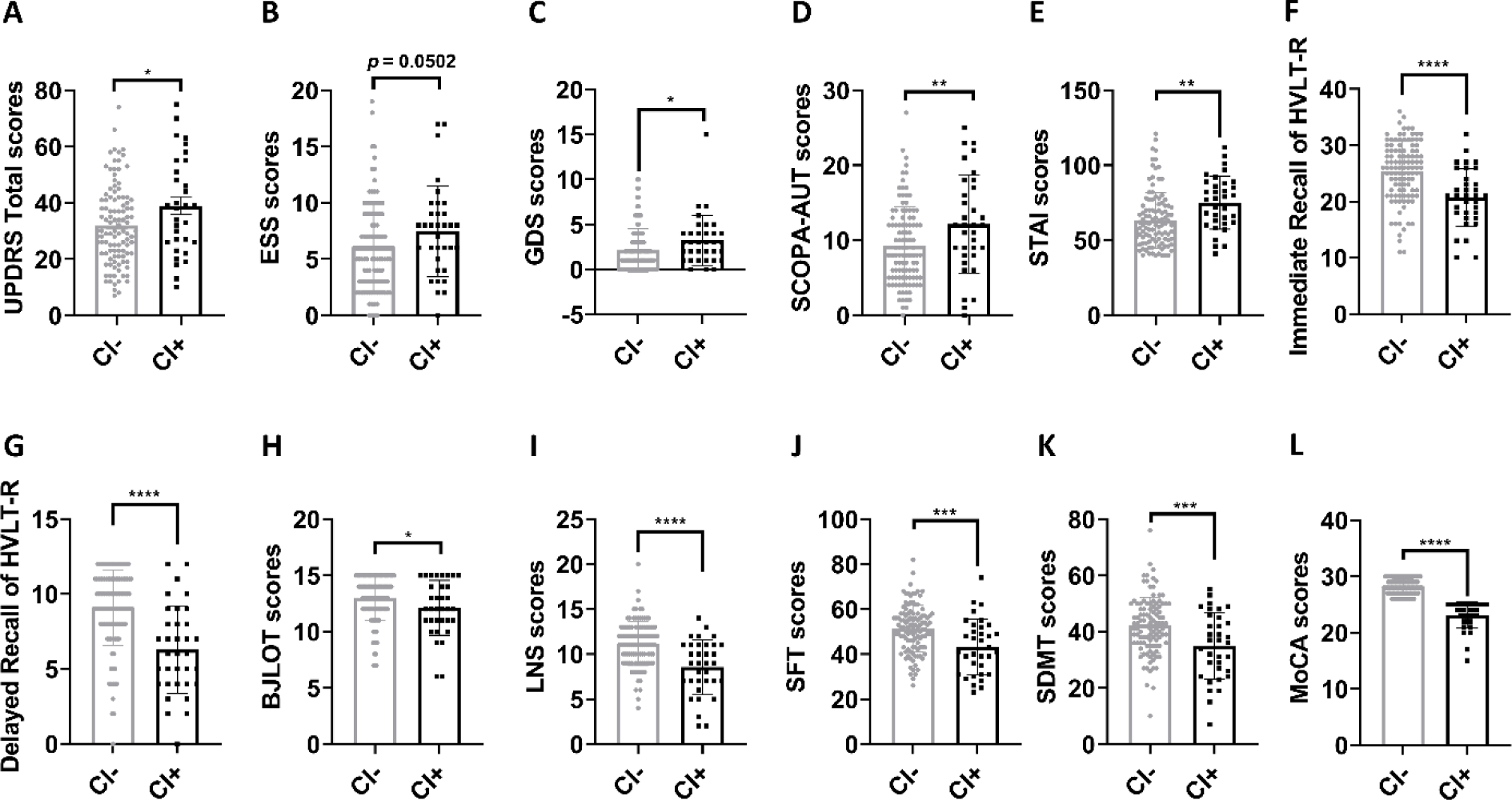
Group differences in clinical characteristics between CI- and CI+ patients. (A-E) CI+ patients exhibited higher scores of total UPDRS (A), ESS (B), GDS (C), SCOPA-AUT (D), and STAI (E) compared to CI-patients. (F-L) CI+ patients showed lower scores of Immediate Recall and Delayed Recall of HVLT-R, BJLOT, LNS, SFT, SDMT, and MoCA compare to CI-patients. The unpaired t-test (CI-group *vs* CI+ group) was used to compare clinical variables. *p* < 0.05 was considered to be statistically significant. \**p* < 0.05, ** *p* < 0.01, \*\*\**p* < 0.001, **** *p* < 0.0001. Abbreviations: CI, cognitive impairment; UPDRS, Unified Parkinson’s Disease Rating Scale; ESS, Epworth Sleepiness Scale; GDS, Geriatric Depression Scale; SCOPA-AUT, Scale for Outcomes in Parkinson’s Disease-Autonomic; STAI, State-Trait Anxiety Inventory; HVLT-R, Hopkins Verbal Learning Test–Revised; BJLOT, Benton Judgment of Line Orientation test; LNS, Letter Number Sequencing test; SFT, Semantic Fluency Test; SDMT, Symbol Digit Modalities Test; MoCA, Montreal Cognitive Assessment.

**Table 1.** Clinical characteristics of CI- and CI+ patients.

| Characteristics | All patients<br>(n = 145) | CI- patients<br>(n = 111) | CI+ patients<br>(n = 34) | Statistic *p* value<br>($\chi^2$ or unpaired t-test) |
| --- | --- | --- | --- | --- |
| Age (years) | 61.15 ± 9.18 | 59.82 ± 9.23 | 65.47 ± 7.64 | **\*\**p* = 0.0015** |
| Sex (Female/Male) | 52/93 | 40/71 | 12/22 | *p* > 0.9999 |
| Education (years) | 15.23 ± 2.97 | 15.42 ± 3.03 | 14.62 ± 2.73 | *p* = 0.1674 |
| Disease duration (years) | 2.04 ± 2.15 | 2.13 ± 2.35 | 1.76 ± 1.33 | *p* = 0.3891 |
| UPDRS-III | 20.82 ± 9.67 | 20.33 ± 9.50 | 22.53 ± 10.20 | *p* = 0.2578 |

Values are mean ± standard deviation. Sex ratio was evaluated with Chi-square test, other characteristics were compared using unpaired t-test. **\*\**p* < 0.01**. Key: CI, Cognitive impairment; UPDRS-III, Unified Parkinson's Disease Rating Scales part III.

### 2.2. Image acquisition

The MRI images were acquired in three dimensions using 3T Siemens scanners (TIM Trio or Verio) from Siemens Healthcare. The scanners utilized a MPRAGE sequence to obtain T1-weighted MRI images. The T1 images were taken with the following settings: Repetition time (TR) = 2300 ms, Echo time (TE) = 2.98 ms, Voxel size = 1 mm^3^, Slice thickness = 1.2 mm, Twofold acceleration, and a sagittal-oblique angulation. The DTI was performed using the following settings: TR = 8,400-8,800 ms, TE = 88 ms, Voxel size = 2 mm^3^, Slice thickness = 2 mm, 64 different directions, and a b-value of 1000 s/mm^2^.

### 2.3. Imaging preprocessing

The DTI images of 145 PD patients were preprocessed using the FMRIB Software Library toolkit (FSL), which can be found at the following website: https://fsl.fmrib.ox.ac.uk/fsl/fslwiki. In short, the initial processes involved correcting head motions, addressing distortions caused by eddy-currents, and mitigating susceptibility artifacts due to inhomogeneity of the magnetic field. Subsequently, DTI metrics including but not limited to fractional anisotropy (FA), mean diffusivity (MD), axial diffusivity, and radial diffusivity were produced. Afterwards, the images of each participant were reconstructed again in the standard MNI space to generate structural network matrix.

### 2.4. Network construction

A free MATLAB toolkit called PANDA, available at http://www.nitrc.org/projects/panda/, was utilized to perform deterministic fiber tractography and create the white matter network. The Fiber Assignment by Continuous Tracking (FACT) algorithm was used to derive the white matter fibers throughout the entire brain connecting 90 nodes in the Automated Anatomical Labelling (AAL) atlas, including both cortical and subcortical areas. The FA skeleton threshold was set to 0.20, and a threshold of 45° was set for the fiber angle. Following the process of white matter tractography, a white matter network matrix was created for each participant. This matrix was based on the fiber number (FN) present in the structural network.

### 2.5. Graph-based network analysis

The brain network measurements were calculated using GRETNA [54], a software that focuses on graphical analysis of networks in the brain. To calculate the global and nodal network measurements, a variety of network density thresholds ranging from 0.05 to 0.50 in steps of 0.05 were used. The area under curve (AUC) was computed for both global and nodal network measurements. The global network metrics included global efficiency, local efficiency, and small-worldness properties, such as clustering coefficient (Cp), characteristic path length (Lp), normalized clustering coefficient (γ), normalized characteristic path length (λ), and small worldness (σ). The nodal network metrics included nodal betweenness centrality (BC), nodal degree centrality (DC), nodal Cp, nodal efficiency, nodal local efficiency, and nodal shortest path length. Previous studies have thoroughly documented the specific definitions for each network measurement [55–57].

### 2.6. Statistical analysis

#### 2.6.1. Comparison of clinical variables

The unpaired t-test (CI-group *vs* CI+ group) was used to compare continuous variables. The χ^2^ test was used to compare variables that were in categories. A *p*-value below 0.05 was considered to be statistically significant.

#### 2.6.2. Comparison of global network strength

Unpaired two sample t-test was conducted to evaluate and compare the overall connectivity strengths of brain networks between CI-group and CI+ group using Network-Based Statistic (NBS) software [58], which can be found at the following link: https://www.nitrc.org/projects/nbs/. A *p*-value below 0.05 after false discovery rate (FDR) correction [59] was considered to be statistically significant. During the NBS analysis, covariates such as age, sex, years of education, and duration of the disease were adjusted.

#### 2.6.3. Comparison of topological network metrics

To compare the global and nodal network measurements, two-way ANOVA test was conducted, and FDR corrections were applied afterwards. A *p*-value lower than 0.05 after FDR correction was considered to have statistical significance. The AUC of global network metric was evaluated by conducting unpaired t-test, and a *p*-value lower than 0.05 was considered to be statistically significant.

#### 2.6.4. Comparison of white matter fiber numbers

To compare the fiber numbers of key nodes, two-way ANOVA test was performed, and FDR corrections were applied afterwards. A *p*-value lower than 0.05 after FDR correction was considered to be statistically significant.

#### 2.6.5. Association analysis

The univariate correlation analysis and multivariate regression analysis with age, sex, disease duration, and years of education as covariates were used to perform association analysis. A *p*-value less than 0.05 was considered statistically significant for associations between MoCA scores and clinical variables. FDR-corrected *p*-value less than 0.05 was considered statistically significant for associations between clinical variables and graphical network metrics.

#### 2.6.6. Mediation analysis

The mediation analysis was carried out using IBM SPSS Statistics Version 26. The age, EDS, and depression were included as the independent variables. The MoCA scores or categorical CI (CI-= 1, CI+ = 2) were enrolled as dependent variables. The topological network metrics that were associated with MoCA scores served as mediators. We simulated the mediation effects of network metrics on the relationships between age, EDS, depression and MoCA scores. In the mediation analysis, confounding variables such as age, sex, disease duration, and years of education were adjusted. Standardized β, t, and *p* values were reported for mediation analysis. A *p*-value less than 0.05 was considered to have statistical significance.

## 3. Results

### 3.1. Group differences in clinical assessments

The demographic and clinical data of CI- and CI+ patients were shown in Table 1 and Figure 1. Compared to CI-patients (n = 111), CI+ patients (n = 34) exhibited older age (*p* = 0.0015, 59.82 ± 9.23 *vs* 65.47 ± 7.64; Table 1), however, the sex distribution, years of education, duration of disease, and motor symptoms measured by UPDRS-III were not significantly different (all *p* > 0.05). Interestingly, CI+ patients showed higher total UPDRS scores (*p* < 0.05; Fig. 1A), ESS scores (*p* = 0.0502; Fig. 1B), GDS scores (*p* < 0.05; Fig. 1C), SCOPA-AUT scores (*p* < 0.01; Fig. 1D), STAI scores (*p* < 0.01; Fig. 1E) and lower scores of Immediate Recall of HVLT-R (*p* < 0.0001; Fig. 1F), Delayed Recall of HVLT-R (*p* < 0.0001; Fig. 1G), BJLOT (*p* < 0.05; Fig. 1H), LNS (*p* < 0.0001; Fig. 1I), SFT (*p* < 0.001; Fig. 1J), SDMT (*p* < 0.001; Fig. 1K) and MoCA (*p* < 0.0001; Fig. 1L).

### 3.2. Associations between MoCA scores and clinical variables

To examine whether the effects of CI on clinical characteristics of PD were independent of confounding variables, multivariate regression analysis was performed with age, sex, years of education, and disease duration as covariates. As shown in Table 2, the MoCA scores were negatively associated with rigidity scores (*p* < 0.05), UPDRS-III scores (*p* = 0.05), UPDRS Total scores (*p* < 0.01), ESS scores (*p* < 0.01), GDS scores (*p* < 0.01), RBDSQ scores (*p* < 0.01), SCOPA-AUT scores (*p* < 0.01), STAI scores (*p* < 0.0001) and positively associated with scores of SFT (*p* < 0.001), SDMT (*p* < 0.0001), LNS (*p* < 0.0001), Immediate Recall (*p* = 0.0001) and Delayed Recall (*p* < 0.0001) of HVLT-R. When categorical CI (CI-= 1, CI+ = 2) entered as dependent variable in the multivariate regression analysis (Table S1), CI was positively associated with GDS scores (*p* < 0.05), STAI scores (*p* < 0.05) and negatively associated with scores of SFT (*p* < 0.05), LNS (*p* = 0.0002), Immediate Recall (*p* = 0.0002) and Delayed Recall (*p* < 0.0001) of HVLT-R. These results suggested that multiple non-motor features, such as depressive and anxious symptoms, were associated with cognitive decline in PD patients.

**Table 2.** Associations between MoCA scores and clinical assessments.

| Clinical assessments |  |  |  |  |  |  |  |  |
| --- | --- | --- | --- | --- | --- | --- | --- | --- |
|  | **HY stage** | **Tremor** | **Rigidity** | **UPDRS-III** | **UPDRS-Total** | **ESS** | **GDS** | **RBDSQ** |
| **$\beta$ value** | $\beta = -0.01$ | $\beta = -0.19$ | $\beta = -0.22$ | $\beta = -0.65$ | $\beta = -1.65$ | $\beta = -0.41$ | $\beta = -0.24$ | $\beta = -0.24$ |
| **$p$-value** | $p = 0.4956$ | $p = 0.0921$ | $p = 0.0278$ | $p = 0.0544$ | $p = 0.0012$ | $p = 0.0013$ | $p = 0.0041$ | $p = 0.0081$ |
| Clinical assessments |  |  |  |  |  |  |  |  |
|  | **SCOPA-AUT** | **STAI** | **BJLOT** | **SFT** | **SDMT** | **LNS** | **HVLT-R**<br>**Immediate Recall** | **HVLT-R**<br>**Delayed Recall** |
| **$\beta$ value** | $\beta = -0.49$ | $\beta = -2.46$ | $\beta = 0.11$ | $\beta = 1.23$ | $\beta = 1.51$ | $\beta = 0.46$ | $\beta = 0.63$ | $\beta = 0.42$ |
| **$p$-value** | $p = 0.0089$ | $p < 0.0001$ | $p = 0.0856$ | $p = 0.0004$ | $p < 0.0001$ | $p < 0.0001$ | $p = 0.0001$ | $p < 0.0001$ |

The data were shown as the $\beta$ and $p$ -values derived from multivariate regression analysis with age, sex, years of education, and disease duration as covariates. The motor function examination was assessed in ON state. $p < 0.05$ was considered statistically significant. Abbreviations: HY, Hoehn & Yahr; UPDRS, Unified Parkinson's Disease Rating Scale; ESS, Epworth Sleepiness Scale; RBDSQ, REM Sleep Behavior Disorder Screening Questionnaire; GDS, Geriatric Depression Scale; SCOPA-AUT, Scale for Outcomes in Parkinson's Disease-Autonomic; SDMT, Symbol Digit Modalities Test; LNS, Letter Number Sequencing; SFT, Semantic Fluency Test Score; STAI, State-Trait Anxiety Inventory; BJLOT, Benton Judgement of Line Orientation; MoCA, Montreal Cognitive Assessment; HVLT-R, Hopkins Verbal Learning Test-Revised.

### 3.3. Group differences of network metrics

The global network metrics, such as global efficiency and local efficiency, were not statistically different between CI- and CI+ patients (FDR-corrected *p* > 0.05). For nodal network metrics, CI+ patients showed lower BC in left calcarine, right superior occipital gyrus, left middle occipital gyrus, left thalamus and higher BC in bilateral putamen, right superior temporal gyrus, and bilateral middle temporal gyrus compared to CI-patients (all FDR-corrected *p* < 0.05; Fig. 2A). Additionally, CI+ patients had lower DC in right superior occipital gyrus and left thalamus compared to CI-patients (FDR-corrected *p* < 0.05; Fig. 2B). Moreover, CI+ patients showed higher nodal Cp in left middle frontal gyrus (FDR-corrected *p* < 0.05; Fig. 2C).

**Fig. 2.**
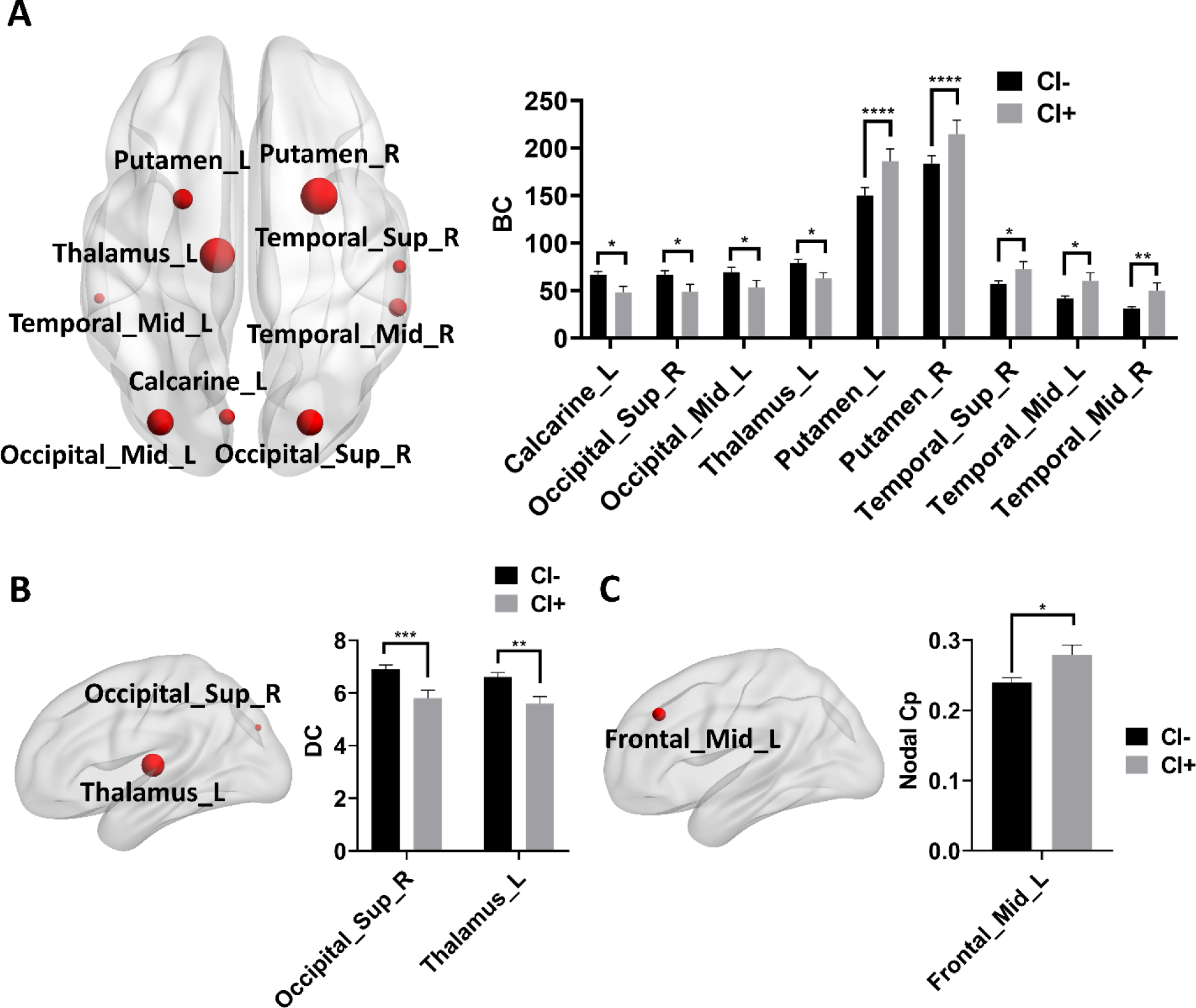
Group differences in structural network metrics between CI- and CI+ patients. (A-C) Group differences in BC (A), DC (B), and nodal Cp (C) between CI- and CI+ patients. Two-way ANOVA test with FDR correction was performed. *p* < 0.05 after FDR correction was considered to have statistical significance. \**p* < 0.05, ** *p* < 0.01, \*\*\**p* < 0.001, **** *p* < 0.0001. Abbreviations: CI, cognitive impairment; BC, Betweenness centrality; DC, Degree centrality; Cp, Clustering coefficient.

### 3.4. Associations between network metrics and MoCA scores

To adjust the effects of potential confounding factors, such as age, sex, education, and disease duration, multivariate regression analysis was performed to examine the associations between MoCA scores and structural network metrics showing statistical difference in Figure 2. We found MoCA scores were significantly associated with nodal BC in left calcarine, right superior occipital gyrus, and right middle temporal gyrus (FDR-corrected *p* < 0.05; Table S2). In addition, MoCA scores were also associated with nodal DC in right superior occipital gyrus and left thalamus and nodal Cp in left middle frontal gyrus (FDR-corrected *p* < 0.05; Table S2).

### 3.5. The effects of age on MoCA scores and structural network metrics

As shown in Table 1, CI+ patients exhibited older age compared to CI-patients. Through association analysis, we also found age was significantly associated with MoCA scores (β = −0.08, *p* = 0.0004) and CI (β = 0.01, *p* = 0.0035), which was independent of sex, years of education, and disease duration. These results suggested that PD patients naturally developed age-dependent cognitive decline. To understand whether age also affected structural network, we analyzed the associations between age and structural networks using univariant Pearson correlation and multivariate regression analysis. As shown in Figure 3, age was significantly associated with BC in right superior occipital gyrus (FDR-corrected *p* < 0.05; Fig. 3A), left putamen (FDR-corrected *p* < 0.05; Fig. 3B), right superior temporal gyrus (FDR-corrected *p* < 0.05; Fig. 3C), right middle temporal gyrus (FDR-corrected *p* < 0.05; Fig. 3D) and DC in right superior occipital gyrus (FDR-corrected *p* < 0.05; Fig. 3E) and left thalamus (FDR-corrected *p* < 0.05; Fig. 3F). Then, we assessed whether structural network metrics played a role in age-dependent cognitive decline in PD patients using mediation analysis. As shown in Figure 4, BC in right superior occipital gyrus (Fig. 4A) and right middle temporal gyrus (Fig. 4B) partially mediated the effects of age on MoCA scores in PD patients. Moreover, DC in right superior occipital gyrus (Fig. 4C) and left thalamus (Fig. 4D) also partially mediated the effects of age on MoCA scores in PD patients. When categorical CI entered as dependent variable in mediation analysis (Fig. S1), we also revealed that BC in right middle temporal gyrus (Fig. S1A) and DC in right superior occipital gyrus (Fig. S1B) and left thalamus (Fig. S1C) partially mediated the effects of age on CI in PD patients.

**Fig. 3.**
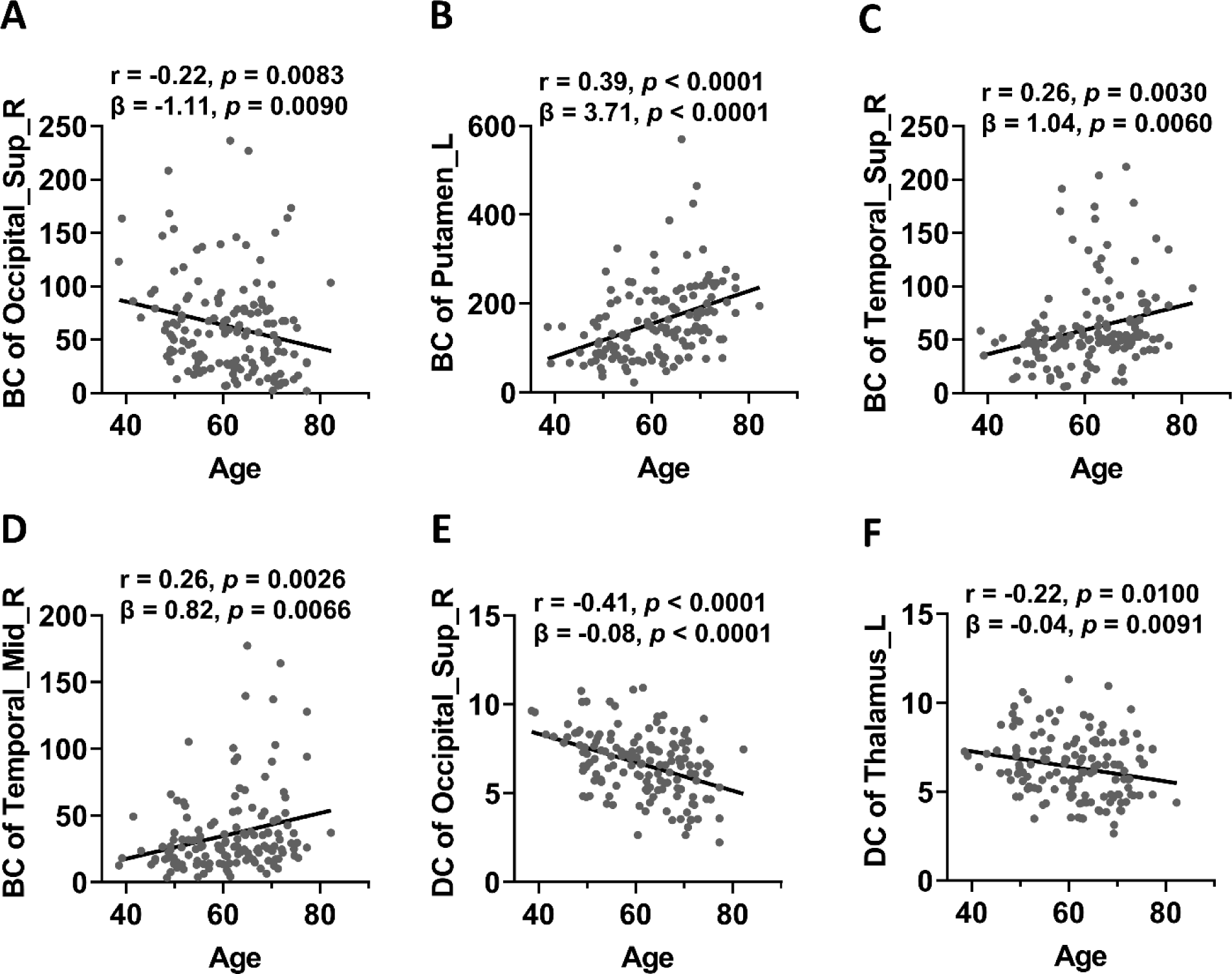
Associations between age and nodal network metrics. (A-D) Age was significantly associated with BC of right superior occipital gyrus, left putamen, right superior temporal gyrus, and right middle temporal gyrus (FDR-corrected *p* < 0.05 in both Pearson correlation analysis and multivariate regression analysis). (E-F) Age was significantly associated with DC of right superior occipital gyrus and left thalamus (FDR-corrected *p* < 0.05 in both Pearson correlation analysis and multivariate regression analysis). The association analysis between graphical network metrics and age was conducted by Pearson correlation method and multivariate regression analysis with age, sex, disease duration, and years of education as covariates. FDR-corrected *p* < 0.05 was considered statistically significant. Abbreviations: BC, Betweenness centrality; DC, Degree centrality.

**Fig. 4.**
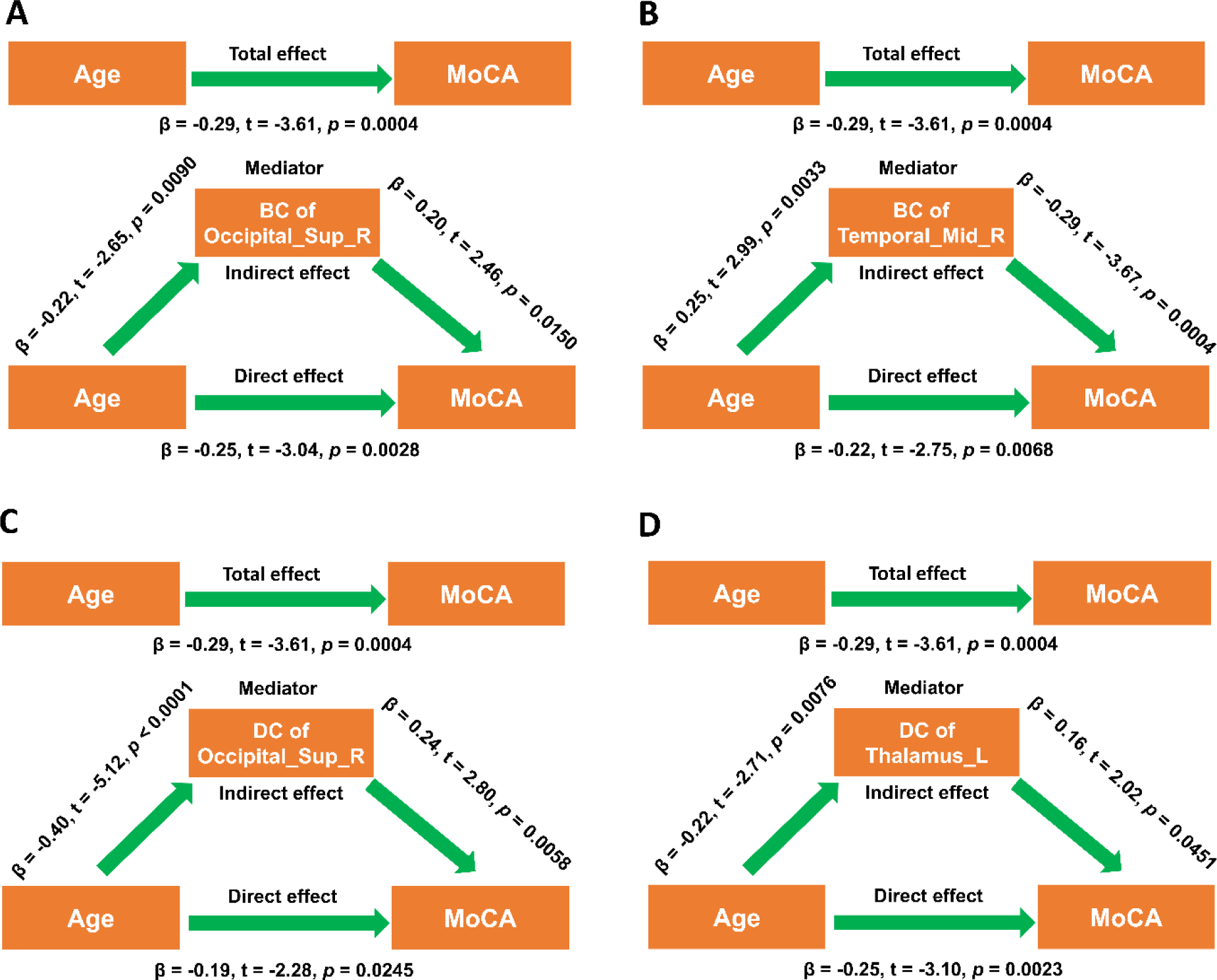
Structural network metrics mediated the effects of age on MoCA scores. (A-B) BC of right superior occipital gyrus and right middle temporal gyrus mediated the effects of age on MoCA scores. (C-D) DC of right superior occipital gyrus and left thalamus mediated the effects of age on MoCA scores. During the mediation analysis, age, sex, disease duration, and years of education were included as covariates. *p* < 0.05 was considered statistically significant. Abbreviations: BC, Betweenness centrality; DC, Degree centrality; MoCA, Montreal Cognitive Assessment.

### 3.6. The effects of EDS on MoCA scores

The Figure 1 showed CI+ patients had higher ESS scores compared to CI-patients (Fig. 1B). In Table 2, MoCA scores were significantly associated with ESS scores. Based on these findings, we hypothesized that EDS measured by ESS may contributed to CI by regulating structural network metrics in PD patients. Initially, ESS scores were significantly associated with DC in right superior occipital gyrus (Fig. S2A). Then, using mediation analysis, we demonstrated that DC in right superior occipital gyrus mediated the negative association between ESS scores and MoCA scores (Fig. S2B). While the mediation analysis of DC in right superior occipital gyrus between ESS scores and CI failed to achieve statistical significance.

### 3.7. The effects of depressive symptoms on MoCA scores

In the Figure 1, CI+ patients had higher GDS scores compared to CI-patients (Fig. 1C). Additionally, MoCA scores were significantly associated with GDS scores (Table 2). Given that depression was associated with cognitive decline in PD and GDS scores were significantly correlated with BC in right middle temporal gyrus (Fig. S3A), we examined whether BC in right middle temporal gyrus mediated the effects of depression on MoCA scores. Actually, we demonstrated that BC in right middle temporal gyrus mediated the effects of depressive symptoms on MoCA scores and cognitive decline (Fig. S3B-C).

### 3.8. The effects of CI on the structural connectivity of key nodes

Because nodal network metrics of right middle temporal gyrus, right superior occipital gyrus, and left thalamus were associated with cognitive decline in PD patients, we examined whether CI shaped structural connectivity of these nodes. As shown in Supplementary Figure S4, the fiber numbers of right middle temporal gyrus, right superior occipital gyrus, and left thalamus were all significantly different between CI- and CI+ patients (Fig. S4A-C).

## 4. Discussion

In this study, we replicated previous findings that global cognition in PD patients was significantly associated with age, EDS, depressive and anxious symptoms, as well as other clinical features. Compared to CI-patients, CI+ patients exhibited more severe non-motor symptoms. In addition, CI+ patients showed dramatical changes of local topological metrics in structural network compared to CI-patients. Furthermore, we demonstrated that different structural topological metrics mediated the negative associations between age, EDS, depression and MoCA scores of PD patients. Based on our findings, we concluded that local topology in structural network was significantly associated with PD-related CI.

### 4.1. The associations between CI and clinical variables

Apart from its typical motor symptoms, such as bradykinesia, static tremor, and rigidity, PD is associated with a heterogeneous non-motor manifestations that remarkably contribute to the overall disease burden. CI is up to 6 times more common in PD patients than in the healthy individuals and is one of the key non-motor features [60]. CI can critically affect quality of life and has been proven to produce substantial economic impacts over the motor symptoms, even at the early stage of PD [61, 62]. According to previous literature, CI was associated with a multitude of clinical features, including age, education, baseline cognition, hypertension, motor symptoms, depressive symptoms, EDS, and anxiety levels [20–24, 63, 64]. In agreement with these results, we observed MoCA scores were significantly associated with age, UPDRS-III scores, ESS scores, GDS scores, and STAI scores (Table 2). Additionally, we also revealed CI was associated with age, GDS scores, and anxiety scores (Table S1). Consistently, CI+ patients showed older age, higher GDS scores, ESS scores, and STAI scores compared to CI-patients (Fig. 1). For education, no statistical association between years of education and MoCA scores (*p* = 0.0796) and CI (*p* = 0.6690). Interestingly, UPDRS-III scores were almost significantly associated with MoCA scores (*p* = 0.0544) but not CI (*p* = 0.4784). Taken together, we concluded that PD-associated CI was associated with multiple clinical variables, especially age, depression, and EDS. Previous studies showed that *APOE* and *SNCA* polymorphisms were associated with CI in PD patients [20, 24], indicating CI in PD may be also modulated by genetic variations. In addition, PD-associated CI was also associated with multiple biochemical variables, such as blood levels of triglyceride and apolipoprotein A1, as well as CSF level of Kininogen-1 [6, 20]. These findings suggested that PD-associated CI may have a complex pathophysiology and was associated with multi-level variables and characteristics.

Age was a major risk factor of PD and associated with both motor and non-motor symptoms [65, 66]. In our recent study, age was significantly associated with scores of LNS, BJLOT, SDMT, SFT, MoCA, and Immediate Recall of HVLT-R of PD patients, which supported that age was a key modifier of cognitive features in PD patients [25]. In current study, we also found age was associated with MoCA scores (β = −0.08, *p* = 0.0004) and CI (β = 0.01, *p* = 0.0035). Therefore, age was a major contributor of CI in PD patients. Indeed, age has been demonstrated to be a key predictor of CI in PD patients [21, 24]. EDS was one of the most prominent non-motor features of PD patients [67–70]. According to a previous meta-analysis, the prevalence of EDS in PD was approximately 35% and EDS was associated with worse motor and non-motor manifestations [70], especially impairment of executive control and processing speed [22, 23]. In our study, we revealed that ESS scores were negatively associated with MoCA scores (*p* = 0.0013), suggesting EDS may contribute to CI in PD patients. Depression had deleteriously detrimental impacts on life quality of PD patients [71]. The incidence of depression in PD was approximately 20-30% [71]. Depression could precede the onset of motor symptoms; therefore, it has been considered as a prodromal PD [72, 73]. According to previous literature, depression is associated with CI in PD patients [74, 75], especially executive dysfunction [76]. In our study, GDS scores were negatively associated with MoCA scores (*p* = 0.0041) and positively associated with CI (*p* = 0.0146), which also supported depression was significantly associated with CI in PD patients. PD patients displayed high anxiety levels than healthy controls [77]. Anxiety was associated with both motor and non-motor symptoms in PD patients [63, 78–80]. Accumulated studies have shown that higher anxiety level was associated with CI of PD patients [63, 64, 78]. Consistently, in our study, we also found MoCA scores and CI were associated with anxiety levels measured by STAI, which suggested anxiety exerted an essential impact on cognitive ability of PD patients. Taken together, age, EDS, depression, and anxiety were important clinical variables tremendously associated with cognitive dysfunction in PD patients.

### 4.2. The key nodes in structural network associated with PD-related CI

Cognitive decline is prevalent in PD patients and significantly associated with both motor and non-motor manifestations. However, the neural mechanisms underlying cognitive decline in PD patients remain largely unclear. fMRI has been used to decode the neural correlates of CI in PD patients and both structural and functional measurements were found to be associated with cognitive dysfunction in PD patients [25, 34, 45]. In our study, local topological metrics of multiple brain nodes were significantly different between CI- and CI+ patients. These nodes distributed in both cortical (temporal, occipital, and frontal lobe) and subcortical regions (bilateral putamen and thalamus) and have been found to be associated with human cognitive function in PD patients [81]. Consistently, most of these nodal metrics also exhibited cognitive level-dependent changes in PD patients [49]. Therefore, PD patients with CI displayed consistently specific patterns of structural network topology compared to PD patients without CI. In this study, nodal BC in left putamen and right putamen was significantly different between CI- and CI+ patients, which suggested putamen function and structure might be associated with cognitive function of PD patients. Actually, CI+ PD patients was found to have reduced putamen volumes compared to controls and putamen volumes showed moderate associations with executive functions [81]. In addition, the functional connectivity between putamen and right cerebellum lobules VI/I was positively correlated with MoCA scores [82]. Furthermore, multiple metabolic pathways in putamen were also significantly different between CI- and CI+ PD patients [83]. Therefore, putamen may be impaired in CI+ PD patients and associated with cognitive decline. We showed BC and DC of left thalamus was significantly reduced in CI+ patients compared to CI-patients, indicating left thalamus was specifically impaired in CI+ patients. In agreement with our findings, left thalamus volume was found to be significantly reduced in CI+ patients [84]. Melief *et al*. (2018) have demonstrated that deficiency of glutamate signaling from thalamus to dorsal striatum induced the impairment of processing speed in PD [85]. Thus, thalamus may be also involved in the impairment of cognitive function in PD patients. We found BC and DC in core nodes of visual system (i.e., right superior occipital gyrus) were significantly different between CI- and CI+ patients, indicating that local network topology in visual network was preferentially impaired in CI+ patients. Actually, previous studies have shown that occipital regions were specifically impaired and associated with cognitive function in PD patients. For example, Chen *et al*. (2019) reported that reduced gray matter volume in the lateral occipital cortex was associated with cognitive dysfunction in PD patients [86]. Additionally, CI+ patients exhibited statistically significant increases of theta band powers in left occipital cortex, which was associated with visuospatial function of PD patients [87]. Furthermore, occipital hypoperfusion was considered to underlie impairment of visual cognition in PD patients without dementia [88]. These results suggested that the impairments of occipital regions may be associated with cognitive decline in PD patients. Additionally, we observed BC in bilateral temporal nodes were significantly increased in CI+ patients, suggested that the structure and function of bilateral temporal lobe was specifically affected in CI+ patients. According to previous studies, CI+ patients exhibited lower gray matter volume in temporal lobe compared to CI-patients [89]. In addition, Chiang *et al*. (2018) revealed that disruption of bilateral mesial temporal lobes in both architecture and functional connectivity contributed to CI in PD [90]. Moreover, the hypometabolism of left middle temporal gyrus was considered to be associated with cognitive deficits in PD patients [91]. Taken together, we concluded that CI-associated key brain nodes in PD distributed in temporal, occipital, and subcortical areas.

### 4.3. Age, structural network, and PD-associated CI

According to previous literature, PD patients exhibited extensive impairment of white matter integrity [92, 93], which was associated with faster disease progression in PD [94]. Additionally, the burden of white matter hyperintensity was a powerful predictor of cognitive decline in PD patients [95–97]. In a recent study, we revealed age-associated changes in white matter network metrics were causally associated with cognitive assessments of PD patients [25]. Specifically, age was negatively associated with hierarchy, global efficiency, local efficiency, small-worldness Cp and positively associated with small-worldness Lp, γ, and σ in structural network [25]. Through mediation analysis, we further demonstrated that global efficiency, local efficiency, and small-worldness Lp of structural network mediated the negative relationship between age and semantic fluency of PD patients [25]. In addition, these global topological measurements also mediated the effects of age on SDMT scores in PD patients [25]. Importantly, age-dependent changes of white matter integrity also contributed to cognitive decline in PD patients. For instance, FA of bilateral inferior cerebellar peduncle partially mediated the effects of age on SDMT scores of PD patients [25]. However, the limitation of our recent study [25] was that no associations between global topological metrics and MoCA scores were observed, implying that local topological metrics may mediate the effects of age on MoCA scores. In current study, we initially revealed age was significantly associated with multiple local topological metrics in structural network, which further demonstrated that ageing is a major contributor of structural network abnormalities in PD [25]. Using mediation analysis, we further demonstrated that age-dependent decline of DC in right superior occipital gyrus and left thalamus contributed to CI in PD. In addition, we also revealed age-dependent increase in BC of right middle temporal gyrus was causally associated with CI in PD. In agreement with these results, recently, we also revealed DC of right superior occipital gyrus and BC of right middle temporal gyrus mediated the negative association between age and cognitive function of PD patients [49]. To summarize, our findings suggested that local topological metrics were associated with age-dependent cognitive decline in PD patients.

### 4.4. EDS, structural network, and PD-associated CI

According to previous literature, EDS was associated with rapid motor progression and severe motor symptoms [22, 67, 98]. In addition, EDS was also linked to more severe non-motor symptoms, including depressive symptoms [68, 69, 99], cognitive decline [23, 100, 101], autonomic dysfunction [69, 99], and sleep disorders [22, 102]. In a recent study, we revealed DC of left calcarine in structural network was negatively associated with ESS scores and DC of left calcarine in structural network mediated the effects of *BIN3* rs2280104 on both ESS scores and EDS [26]. Therefore, local topological metrics were associated with EDS of PD patients. Consistently, in current study, we revealed ESS scores were significantly associated with DC of right superior occipital gyrus. Using mediation analysis, we further revealed that DC of right superior occipital gyrus mediated the negative association between ESS scores and MoCA scores. Interestingly, the significant association between DC of right superior occipital gyrus and cognitive function of PD patients has been revealed in a recent study [49]. These results suggested that reduction of DC in right superior occipital gyrus during aging also contributed to EDS-associated cognitive decline.

### 4.5. Depression, structural network, and PD-associated CI

It has been shown that depressed PD patients showed significant changes in connecting white matter tracts among prefrontal-temporal regions compared to non-depressed PD patients and healthy controls [103]. In another study, depression scores were found to be associated with disruptions of white matter tracts connected to thalamic subnuclei [104]. Moreover, it seemed that the global efficiency and small-worldness Lp of the structural brain network were impaired in PD patients with depression [105]. These findings suggested that structural network impairments contributed to depression in PD patients. In current study, we revealed BC of right middle temporal gyrus in structural network was higher in CI+ patients than CI-patients, which was consistent with our recent study showing PD patients with lower SDMT scores exhibited greater BC of right middle temporal gyrus than PD patients with higher SDMT scores [49]. Here, we demonstrated that BC of right middle temporal gyrus mediated the effects of depressive symptoms on MoCA scores and CI of PD patients, suggesting higher BC of right middle temporal gyrus in PD was causally correlated with PD-related CI. Importantly, the significant association between BC of right middle temporal gyrus and cognitive function of PD patients has been confirmed in our recent study [49]. Taken together, our findings suggested that depression contributed to CI in PD via the regulation of local topological metrics in right middle temporal gyrus.

### 4.6. Strengths and limitations of this study

In this study, we replicated previous findings that age, EDS, and depressive symptoms were significantly associated with cognitive decline of PD patients. Using network analysis and mediation analysis, we demonstrated that different local topological metrics mediated the effects of age, EDS, and depression on global cognition of PD patients. Therefore, a major contributor of our study to the filed was that we provided network-level explanations to the associations between age, EDS, depression and cognitive decline in PD patients. Importantly, we identified some key nodes, such as right middle temporal gyrus and right superior occipital gyrus, were network mediators of cognitive decline in PD patients, which have been confirmed in a recent study [49]. Another interesting finding was that local topological metrics in white matter network greatly contributed to age-dependent cognitive decline in PD patients, which was consistent with our recent findings [49]. Recently, we have demonstrated that global network metrics contributed to semantic fluency and global cognition of PD patients [25]. Therefore, our findings expanded present understating of the network topological correlates mediating the effects of age on global cognition of PD patients. The limitation of this study was its cross-sectional design; thus, our findings were required to be validated in future longitudinal studies.

## 5. Conclusions

CI+ PD patients exhibited worse non-motor symptoms than CI-PD patients. Age, EDS, depression and anxiety were negatively associated with global cognition of PD patients. CI+ PD patients exhibited statistically different local topological properties in structural network compared to CI-PD patients. Different local topological metrics mediated the negative associations between age, EDS, depression and global cognition of PD patients.

## Author contributions

Zhichun Chen, Conceptualization, Formal analysis, Visualization, Methodology, Writing, review and editing; Guanglu Li, Data curation, Formal analysis, Visualization; Liche Zhou, Data curation, Formal analysis, Investigation; Lina Zhang, Formal analysis, Investigation, Methodology; Li Yang, Supervision, Writing, review and editing; Jun Liu, Conceptualization, Supervision, Funding acquisition, Writing, review, and editing.

## Acknowledgments

Data used in the preparation of this article were obtained from the Parkinson’s Progression Markers Initiative (PPMI) database (www.ppmiinfo.org/data). We thank the share of PPMI data by all the PPMI study investigators. PPMI – a public-private partnership – is funded by the Michael J. Fox Foundation for Parkinson’s Research and funding partners, which can be found at www.ppmiinfo.org/fundingpartners.

## Data availability

All the raw data used in the preparation of this Article were downloaded from PPMI database (www.ppmi-info.org/data).All data produced in the present study are available upon reasonable request to the authors.

## Supporting Information

Additional supporting information may be found online in the Supporting Information section at the end of the article.

