## Supplemental Files for "Structural network topology mediated cognitive impairment in Parkinson’s disease"

**Supplementary materials**

**Table S1** Associations between CI and clinical assessments.

| Clinical assessments | | | | | | | | | | | | | | | | |
| --- | --- | --- | --- | --- | --- | --- | --- | --- | --- | --- | --- | --- | --- | --- | --- | --- |
|  | | **HY stage** | | **Tremor** | | **Rigidity** | | **UPDRS-III** | | **UPDRS-Total** | | **ESS** | | **GDS** | | **RBDSQ** |
| **β value**  ***p*-value** | | β = 0.07  *p* = 0.4841 | | β = 0.36  *p =* 0.6017 | | β = 0.85  *p* = 0.1571 | | β = 1.43  *p* = 0.4784 | | β = 5.40  *p* = 0.0814 | | β = 1.02  *p* = 0.1805 | | β = 1.21  *p* = 0.0146 | | β = 0.65  *p* = 0.2228 |
| **Clinical assessments** | | | | | | | | | | | | | | | | |
|  | **SCOPA-AUT** | | **STAI** | | **BJLOT** | | **SFT** | | **SDMT** | | **LNS** | | **HVLT-R**  **Immediate Recall** | | **HVLT-R**  **Delayed Recall** | |
| **β value**  ***p*-value** | β = 1.82  *p* = 0.0966 | | β = 9.18  *p* = 0.0120 | | β = -0.33  *p* = 0.3750 | | β = -4.59  *p* = 0.0272 | | β = -3.38  *p* = 0.0705 | | β = -1.76  *p* = 0.0002 | | β = -3.58  *p* = 0.0002 | | β = -2.33  *p* < 0.0001 | |

The data were shown as the β and *p*-values derived from multivariate regression analysis with age, sex, years of education, and disease duration as covariates. The motor function examination was assessed in ON state. *p* < 0.05 was considered statistically significant. Abbreviations: CI, Cognitive impairment; HY, Hoehn & Yahr; UPDRS, Unified Parkinson’s Disease Rating Scale; ESS, Epworth Sleepiness Scale; RBDSQ, REM Sleep Behavior Disorder Screening Questionnaire; GDS, Geriatric Depression Scale; SCOPA-AUT, Scale for Outcomes in Parkinson's Disease-Autonomic; SDMT, Symbol Digit Modalities Test; LNS, Letter Number Sequencing; SFT, Semantic Fluency Test Score; STAI, State-Trait Anxiety Inventory; BJLOT, Benton Judgement of Line Orientation; MoCA, Montreal Cognitive Assessment; HVLT-R, Hopkins Verbal Learning Test-Revised.

**Table S2** The associations between MoCA scores and structural network metrics.

| Nodal BC | | | | | | | | | | | |
| --- | --- | --- | --- | --- | --- | --- | --- | --- | --- | --- | --- |
| **Metrics**  **Statistics** | Calcarine_L | | Occipital_Sup_R | Occipital_Mid_L | Putamen_L | Putamen_R | | Thalamus_L | Temporal_Sup_R | Temporal_Mid_L | Temporal_Mid_R |
| **β value**  ***p*-value** | **β = 3.31**  ****p* = 0.0351** | | **β = 3.66**  ****p* = 0.0450** | β = 1.64  *p* = 0.3843 | β = -4.38  *p* = 0.1517 | β = -1.77  *p* = 0.5215 | | β = 2.62  *p* = 0.1267 | β = -1.75  *p* = 0.2251 | β = -2.14  *p* = 0.1343 | **β = -3.48**  *****p* = 0.0036** |
|  | | **Nodal DC** | | | | | | | | **Nodal Cp** | |
| **Metrics**  **Statistics** | | Occipital_Sup_R | | | | | Thalamus_L | | | Frontal_Mid_L | |
| **β value**  ***p*-value** | | **β = 0.15**  ****p* = 0.0116** | | | | | **β = 0.11**  ****p* = 0.0451** | | | **β = -0.006**  *****p* = 0.0096** | |

Multivariate regression analysis was performed by adjusting age, sex, disease duration, and years of education. False discovery rate (FDR) was used for multiple comparison corrections. FDR-corrected *p* < 0.05 was considered statistically significant. The bold values indicate statistical significance. **p* < 0.05, ***p* < 0.01.

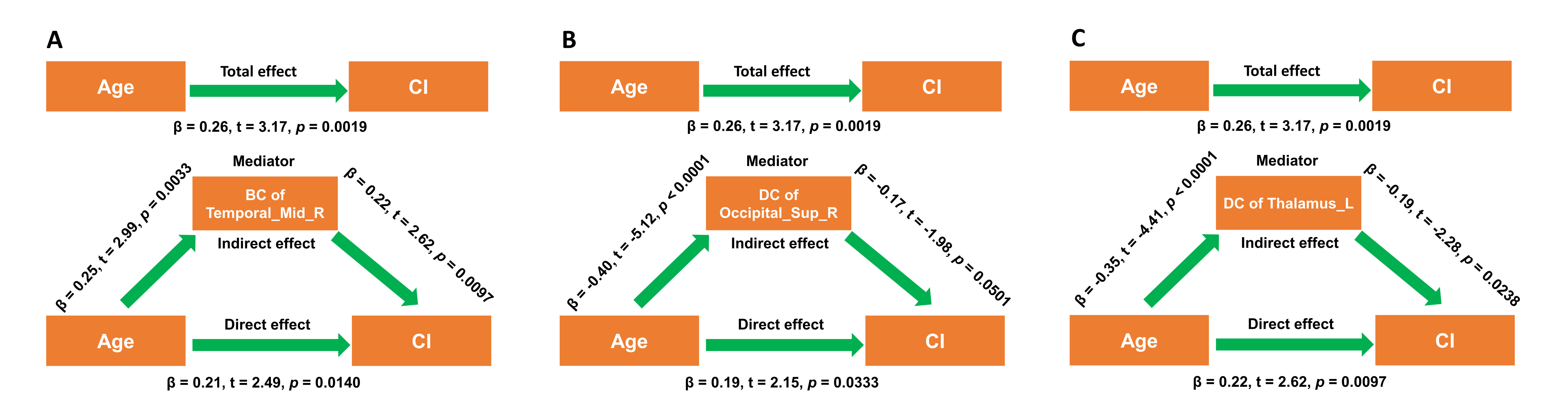

**Supplementary Fig. 1.** **Structural network metrics mediated the effects of age on CI.** (A) BC of right middle temporal gyrus mediated the effects of age on CI. (B-C) DC of right superior occipital gyrus and left thalamus mediated the effects of age on CI. During the mediation analysis, age, sex, disease duration, and years of education were included as covariates. *p* < 0.05 was considered statistically significant. Abbreviations: BC, Betweenness centrality; DC, Degree centrality; CI, Cognitive impairment.

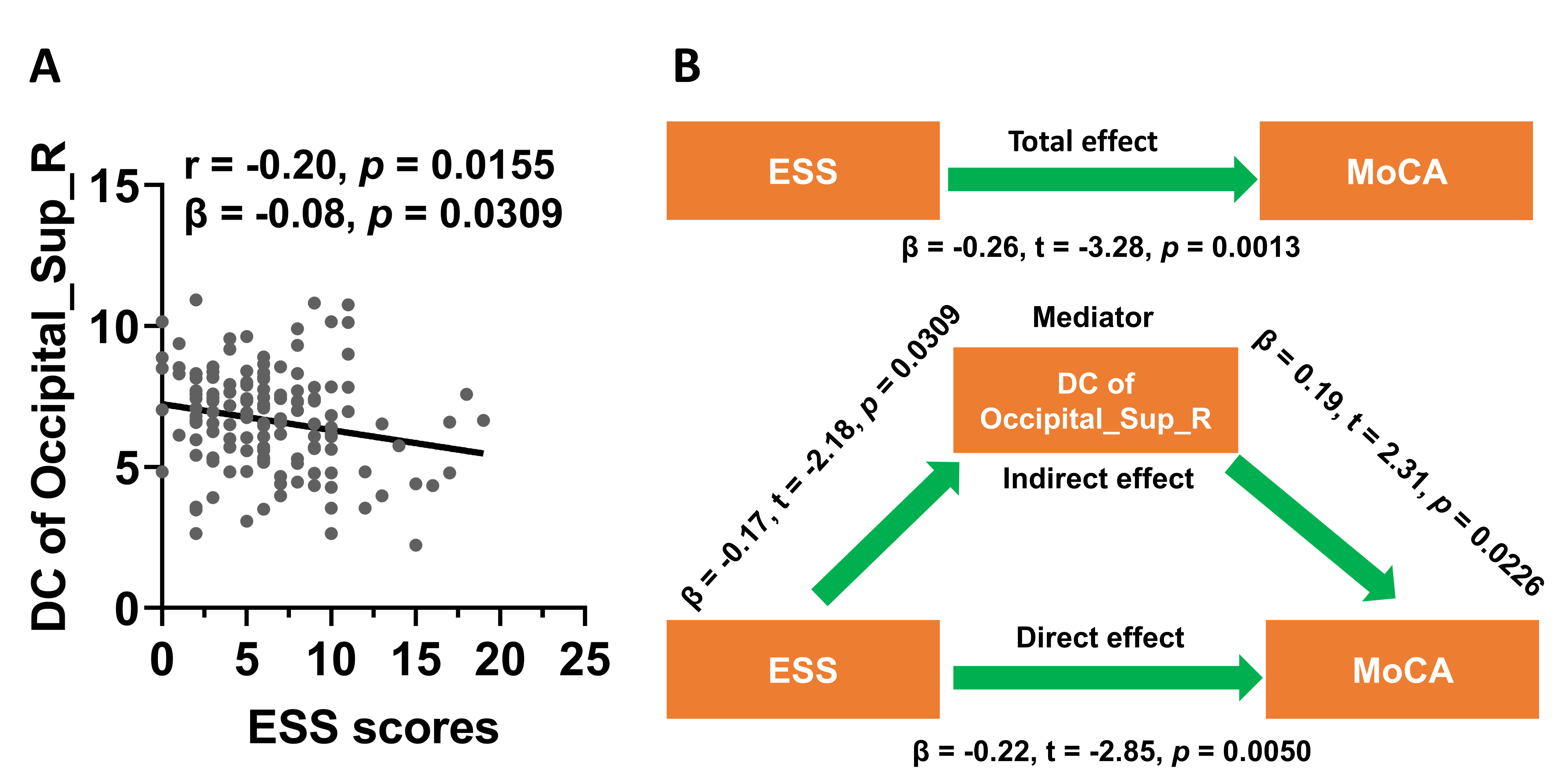

**Supplementary Fig. 2.** **Structural network metrics mediated the negative association between ESS scores and MoCA scores.** (A) ESS scores were negatively associated with DC of right superior occipital gyrus. (B) DC of right superior occipital gyrus mediated the negative association between ESS scores and MoCA scores. The association analysis between graphical network metrics and ESS scores was conducted by Pearson correlation method and multivariate regression analysis with age, sex, disease duration, and years of education as covariates. During the mediation analysis, age, sex, disease duration, and years of education were included as covariates. *p* < 0.05 was considered statistically significant. Abbreviations: DC, Degree centrality; ESS, Epworth Sleepiness Scale; MoCA, Montreal Cognitive Assessment.

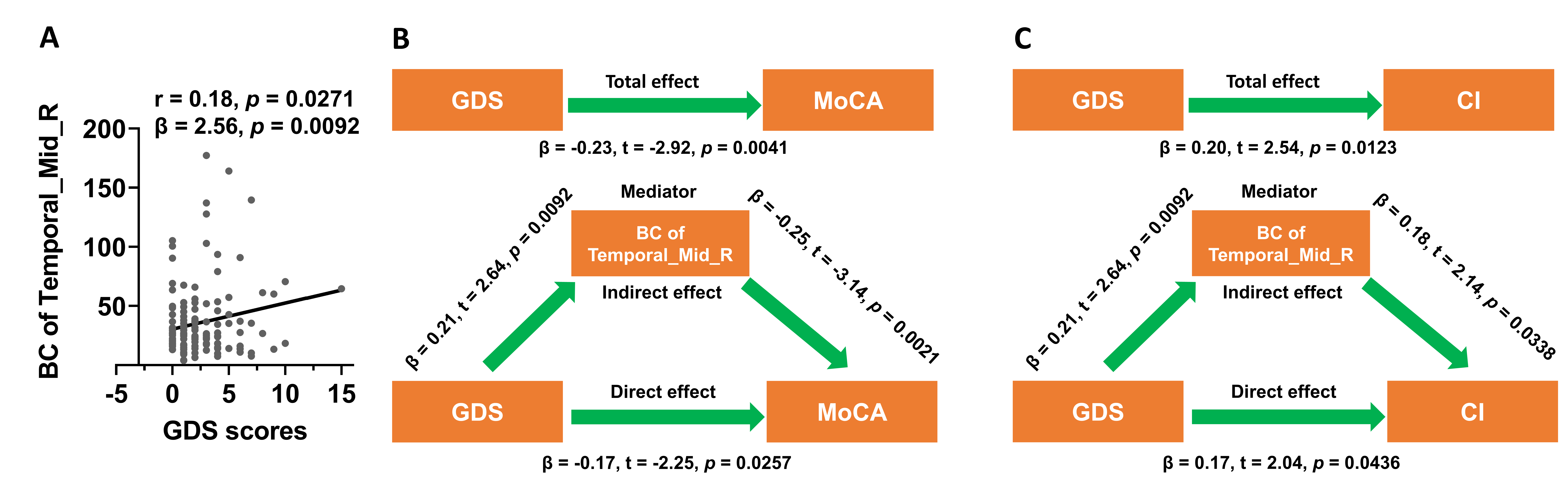

**Supplementary Fig. 3.** **Structural network metrics mediated the effects of depressive symptoms on MoCA scores and cognitive decline.** (A) GDS scores were positively associated with BC of right middle temporal gyrus. (B-C) BC of right middle temporal gyrus mediated the negative association between GDS scores and MoCA scores or CI. The association analysis between graphical network metrics and GDS scores was conducted by Pearson correlation method and multivariate regression analysis with age, sex, disease duration, and years of education as covariates. During the mediation analysis, age, sex, disease duration, and years of education were included as covariates. *p* < 0.05 was considered statistically significant. Abbreviations: CI, Cognitive impairment; BC, Betweenness centrality; GDS, Geriatric Depression Scale; MoCA, Montreal Cognitive Assessment.

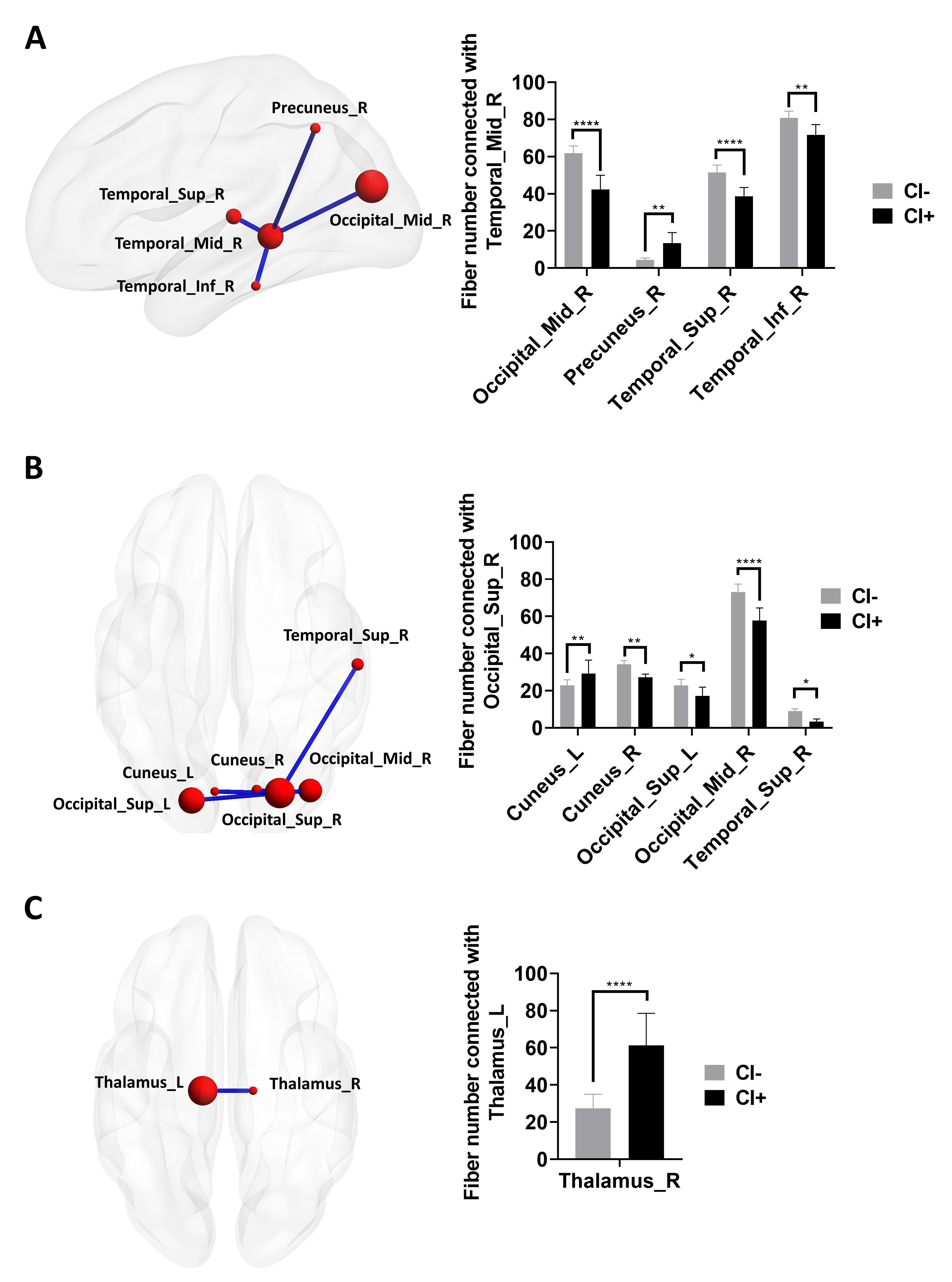

**Supplementary Fig. 4. Group differences in the fiber numbers of key nodes.** (A) Right middle temporal gyrus showed statistically different fiber numbers between CI- and CI+ patients (FDR-corrected *p* < 0.05). (B) Right superior occipital gyrus showed statistically different fiber numbers between CI- and CI+ patients (FDR-corrected *p* < 0.05). (C) The fiber numbers between left thalamus and right thalamus were significantly different between CI- and CI+ patients (FDR-corrected *p* < 0.05). Two-way ANOVA test with FDR correction was performed. *p* < 0.05 after FDR correction was considered to have statistical significance. **p* < 0.05, ** *p* < 0.01, **** *p* < 0.0001. Abbreviations: CI, cognitive impairment.
